# Meningioma Hyperostotic Subtype Defines a TRAF7-Associated Phenotype

**DOI:** 10.64898/2026.01.15.26344124

**Authors:** Aymen Kabir, Abraham Dada, Wesley Shoap, Rithvik Ramesh, Daniel Quintana, Michael A. Torres-Espinosa, Christian Jimenez, Robert C Osorio, Kanish Mirchia, Charlotte D. Eaton, David R. Raleigh, Ezequiel Goldschmidt

**Affiliations:** Department of Neurological Surgery, University of California, San Francisco, San Francisco, CA; Department of Neurosurgery, Louisiana State University, New Orleans, LA; Department of Pathology, University of California, San Francisco, San Francisco, CA; Department of Radiation Oncology, University of California, San Francisco, San Francisco, CA; Institute of Cancer and Genomic Sciences, University of Birmingham, United Kingdom

## Abstract

**Background:** Meningioma-induced hyperostosis (MIH) is a frequent radiographic finding, yet its underlying mechanisms remain poorly understoodWhile hyperostosis has traditionally been treated as a binary phenomenon, the aim of this study was to determine whether MIH represents a heterogenous process with distinct radiological subtypes associated with genetic associations.

**Methods:** We retrospectively reviewed the records and imaging of patients with meningiomas resected between 2021-2024 at a single institution. Somatic mutations identified through next-generation sequencing were analyzed. CT images were analyzed for bone involvement and hyperostosis subtype. Type I hyperostosis was defined by destruction of cortical architecture while Type II hyperostosis was defined by the preservation of cortical structure. Associations with *TRAF7* mutations were assessed using univariate testing, multivariable logistic regression, and supervised machine-learning models. Quantitative bone density analysis was performed using region-of-interest grayscale histogram analysis.

**Results:** Among 384 tumors, 54 (14.1%) exhibited hyperostosis—23 Type I and 31 Type II. *TRAF7* mutations were significantly enriched in Type I hyperostosis compared with Type II and non-hyperostotic tumors (78.3% vs 25.8% vs 17.0%, p<0.001). Type I hyperostosis independently predicted *TRAF7* mutations (OR:18.73, p=0.001), along with skull base location, smaller tumor size, homogeneous contrast enhancement, and extensive T2 hyperintensity. Gradient boosting achieved the highest predictive accuracy (AUC=0.854). Quantitative bone density analysis demonstrated preserved cortical–cancellous architecture in Type II hyperostosis, whereas Type I showed architectural disruption.

**Conclusions:** MIH is a radiographically heterogenous phenomenon. Hyperostosis with disrupted cortical architecture is strongly associated with *TRAF7* mutations and may represent a key feature of this mutation’s radiographic phenotype.

## Introduction

Meningiomas are the only primary intracranial tumor that can induce abnormal growth of the adjacent bone, a term coined meningioma-induced hyperostosis (MIH). Hyperostosis is traditionally defined as hyperplasia of bone adjacent to the tumor and there is wide variability in frequency at which this occurs in patients with meningiomas. Hyperostosis is often the primary cause for clinical presentation and impairs complete tumor removal.^1–3^ MIH is more commonly observed in the skull base and traditionally related to slow tumor growth.^2,5^ Despite developments in meningioma research and classification tools, the exact underlying mechanisms of MIH remain unclear.^4,5^

Recent advances in molecular profiling have shed light on somatic mutations across meningioma subtypes, highlighting various mutations that correlate with specific tumor behaviors, histopathology, and location.^6,7^ Within meningiomas, the most prevalent non-*NF2* mutation is *TRAF7*, found in around 25% of all tumors.^8,9^ Notably, emerging evidence suggests a strong association between *TRAF7* and hyperostosis, particularly with tumors located in the skull base.^10–12^ These findings suggest a potential role of *TRAF7* in bone remodeling processes, although the molecular mechanisms related to these findings remain poorly understood.

Whether MIH is a heterogenous phenomenon, with distinct radiologic and genetic signatures, has not been investigated. In this study, we define two distinct subtypes of MIH, characterized by their differential effects on bony architecture. By comparing these subtypes, we were able to define genetic and radiological factors that could allow us to predict DNA somatic mutations in hyperostotic meningiomas.

## Methods

### Study Population

Patients undergoing surgical resection of meningiomas from 2021 to 2024 at a single institution were identified. Patients with available next-generation sequencing panels and preoperative imaging studies, including CT and MRI, were included in the final analysis. Patients without sufficient imaging studies or those with previous operations that disrupted cortical architecture were excluded from this study. The study was approved by the Ethics Committee.

### Data Collection

Electronic health records of patients included in this study were retrospectively reviewed for patient demographics, tumor characteristics, pathological data, and associated genetic data. Demographic data included patient age and sex. Tumor characteristics collected included tumor location (skull base vs convexity), maximal intradural tumor dimension, and various imaging characteristics seen on MRI and CT. Pathological data collected included pathologically defined WHO grade. Lastly, DNA extracted from surgical samples and examined through capture-based next-generation sequencing panels that target over 500 cancer-related genes at the UCSF Clinical Cancer Genomics Lab highlighted specific somatic mutations that were examined in our study.

### Imaging Characteristics and Hyperostosis

Patient MRI and CT findings were individually reviewed by the senior author, blinded to genetic data at the time of documentation of imaging findings. MRI features assessed included contrast enhancement pattern (homogenous vs heterogenous) and the presence of peritumoral edema, identified by hyperintensity on T2-weighted images. CT features assessed included bone involvement (osteolytic vs hyperostotic) and further characterized by type of hyperostosis. Hyperostosis was defined into two morphological subtypes (Figure 1):

1. Type I: hyperostosis with abnormal cortical-cancellous architecture.
2. Type II hyperostosis with preservation of native bone structure.

**Figure 1.**
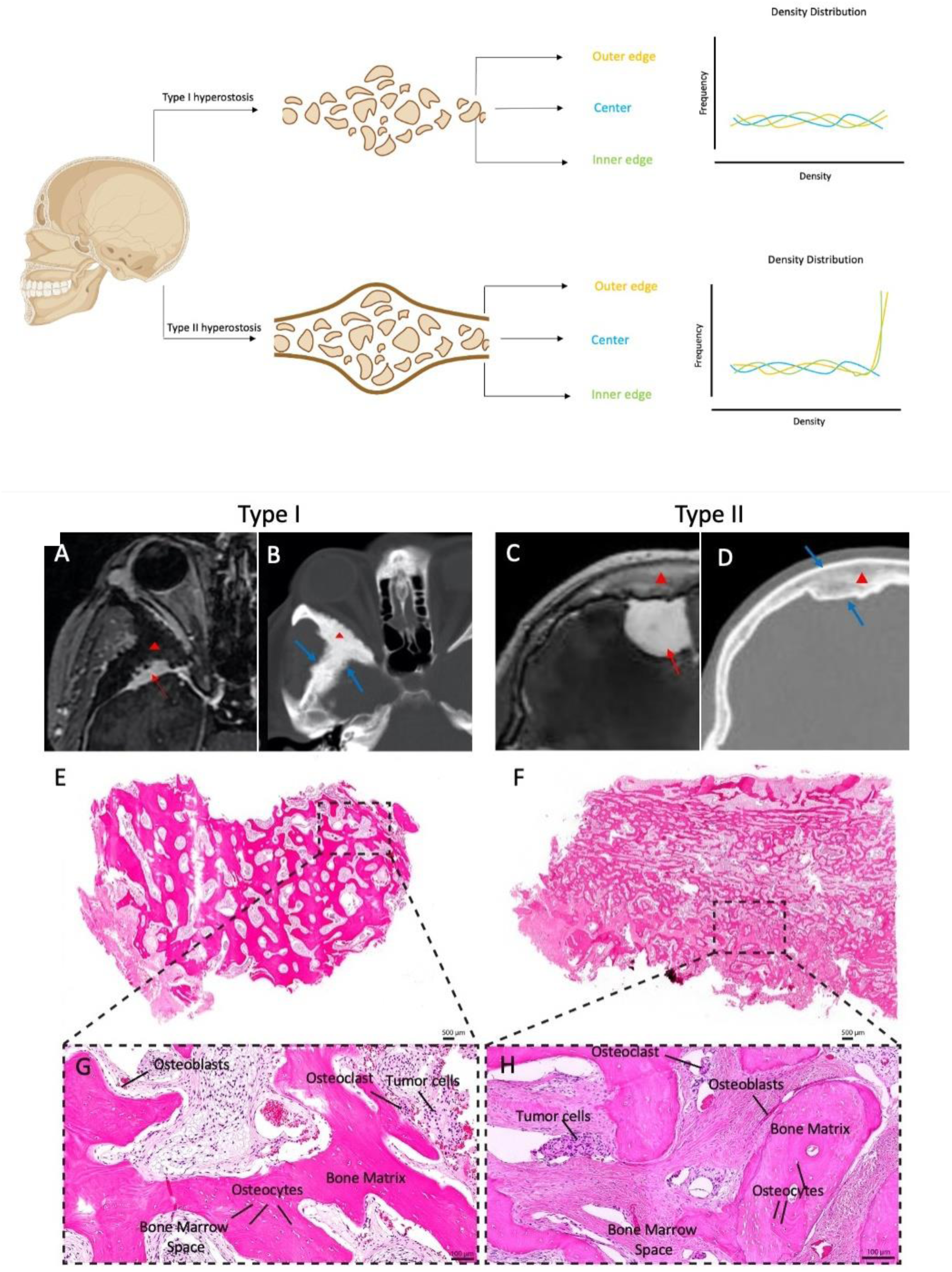
Type I and II Hyperostosis. *Top:* Grayscale histograms from CT-derived ROIs illustrate bone density patterns across the outer edge, center, and inner edge of hyperostosis. In Type I (top), curves overlap with minimal shift, reflecting preserved architecture. In Type II (bottom), outer and inner edge curves show a rightward shift, indicating increased density and structural disruption. *Bottom:* (A) and (B) are a contrasted MR and non-contrasted CT scan depicting a type I hyperostotic sphenoid wing meningioma harboring a *TRAF7* single nucleotide variant. (C) and (D) are the correlates on a parasagittal meningioma with an *NF2* deletion. (E) and (F) and (G) and (H) are low- and high-magnification histological images for both types. The bone marrow spaces are infiltrated with tumor cells; in type I there is relative preservation of the fatty tissue whereas in type II the marrow is replaced with a fibrotic reaction. The intradural tumor is marked with a red arrow, the hyperostotic portion with a red arrowhead. The blue arrows mark the edge of the bony reaction highlighting the lack of cortical-cancellous differentiation in type I but its presence in type II.

### Quantitative Bone Density Analysis

Quantitative bone density analysis was performed using ImageJ. Regions of interest (ROIs) were manually placed within the hyperostotic bone at three standardized locations: the outer cortical edge adjacent to the extracranial surface, a central region at the midpoint between the outer and inner bony edges, and the inner cortical edge adjacent to the intracranial surface.

ROIs were drawn on axial CT slices at the level of maximal hyperostotic involvement. Grayscale intensity histograms were extracted and normalized to account for differences in ROI size. For each group (normal bone, Type I hyperostosis, and Type II hyperostosis), histograms were averaged, and the resulting patterns were compared to evaluate whether the distinction between cortical and cancellous bone was preserved or disrupted.

### Statistical Analysis

Our initial analysis compared the different features collected (radiographic, demographic, molecular, etc.) across three different patient groups defined by: (1) no hyperostosis, (2) Type I hyperostosis, and (3) Type II hyperostosis. Continuous variables were assessed and compared across the three groups using ANOVA. Chi-square tests of Fisher’s exact tests were used when analyzing categorical variables.

Patients were then stratified based on the presence or absence of a *TRAF7* mutation. Continuous variables were analyzed via independent t-tests while categorical variables were assessed using chi-square or Fisher’s exact tests. A multivariable logistic regression model was then used to identify patient- and tumor-specific variables associated with *TRAF7* mutation status. Covariates in this model were selected based on significance in the prior univariate analysis.

To explore non-parametric relationships and identify the predictive value of collected features, we employed various supervised machine learning models including Random Forest, Gradient Boosting Machines (GBM), and Support Vector Machine (SVM). Each model was trained and used to identify the most predictive variables associated with *TRAF7* mutations.

To visualize the distribution of imaging and clinical features we performed principal component analysis on all variables collected for each tumor. Radiographic covariates (tumor location, maximal diameter, sinus invasion, enhancement pattern, cystic change, bone involvement variables, T2 signal characteristics, etc.) were merged with targeted next-generation sequencing results. Each mutation detected at least once was converted to a binary indicator (e.g., MT_TRAF7, MT_NF2) via one-hot encoding, yielding a joint imaging + genomic matrix. Continuous variables were centered and scaled; all others were kept as factors. Rows with missing data were listwise deleted (final n = 384). Pairwise dissimilarities were computed with the Gower metric (daisy, cluster v2.1.5) to accommodate mixed data types. The resulting distance matrix was subjected to classical multidimensional scaling (cmdscale) to obtain a two-dimensional configuration. Ward’s minimum-variance hierarchical clustering (hclust, method = “ward.D2”) was applied to the same distance matrix, and a cut height producing two clusters was chosen a priori. The MDS coordinates were visualized with ggplot2 v3.5.1; points were colored either by cluster assignment or, post-hoc, by TRAF7 mutation status (wild-type = grey, mutant = blue). All analyses were performed in R v 4.4.2 with the tidyverse v2.0.0.

## Results

### Hyperostosis

Our final cohort consists of 384 meningiomas with 54 (14.1%) exhibiting hyperostosis (Table 1). Of those with hyperostosis, 23 (6.0%) exhibited type I hyperostosis with the remaining (n=31, 8.1%) exhibiting what was defined as type II hyperostosis. Figure 1 shows representative histopathological slides of the two defined subtypes. Statistically significant differences between the groups were observed in age, sex, extent of bone involvement, and specific genetic mutations.

**Table 1.** Cohort Characteristics.

| Table 1.<br>Cohort Characteristics |  |  |  |  |
| --- | --- | --- | --- | --- |
| Characteristic | No Hyperostosis | Type I Hyperostosis | Type II Hyperostosis | p Value |
| No. of patients, N=384 | 330 | 23 | 31 |  |
| Age in yrs., mean ± SD | 61.3 ± 15.0 | 50.7±12.4 | 54.0±14.2 | **<0.001\*** |
| Female n (%) | 217 (65.8) | 21 (91.2) | 20 (64.5) | **0.039‡** |
| Location, n (%) |  |  |  | 0.059‡ |
| Convexity | 135 (40.9) | 5 (21.7) | 8 (25.8) |  |
| Skull Base | 195 (59.1) | 18 (78.3) | 23 (74.2) |  |
| MRI Features |  |  |  |  |
| Maximal Tumor Dimension, mean ± SD, N= 372 | 3.70±1.69 | 4.31± 1.76 | 4.24±1.97 | 0.082* |
| Sinus Invasion, n (%), N=321 | 72/267 (27.0) | 6/23 (26.1) | 7/31 (22.6) | 0.871‡ |
| Homogenous Enhancement, n (%) | 231/330 (70.0) | 19/23 (82.6) | 22/31 (71.0) | 0.437‡ |
| Cystic, n (%) | 5/330 (1.5) | 0/23 (0) | 2/31 (6.5) | 0.159† |
| T2 Signal Change, n (%) | 171/330 (51.8) | 10/23 (43.5) | 17/31 (54.8) | 0.720‡ |
| T2 Signal Change 2x Tumor Volume, n (%) N=198 | 17/171 (9.9) | 1/10 (10) | 2/17 (11.8) | 0.972‡ |
| Bone>Tumor | 0/330 (0.0) | 22/23 (95.7) | 0/31 (0.0) | **<0.001‡** |
| Genetic Mutations |  |  |  |  |
| *TRAF7* | 56/330 (17.0) | 18/23 (78.3) | 8/31 (25.8) | **<0.001‡** |
| *NF2* | 140/330 (42.4) | 4/23 (17.4) | 10/31 (32.3) | 0.039‡ |
| *AKT1* | 35/330 (10.6) | 2/23 (8.7) | 5/31 (16.1) | 0.590† |
| *KLF4* | 21/330 (6.4) | 4/23 (17.4) | 1/31 (3.2) | 0.105† |

\* Anova.
† Fisher exact test.
‡ Chi-square test.

Patients with hyperostosis, independent of the type, were younger at presentation than those without bone involvement (mean age: 50.7 ± 12.4 vs. 54.0 ± 14.2 vs. 61.3 ± 15.0 years, p < 0.001). The proportion of female patients was higher in the type I hyperostosis group compared to those without bone involvement and those with type II hyperostosis (91.2% vs. 65.8% vs. 64.5%, p = 0.039). A greater extent of bone involvement, defined by the bone component being larger in size than the tumor, was more frequent in type I hyperostosis than in cases with type II hyperostosis (95.7% vs. 0.0%, p < 0.001).

With respect to genetic alterations, we analyzed potential associations with the most prevalent mutations noted in Table 1. *TRAF7* mutations occurred at significantly higher rates in type I hyperostotic meningiomas (78.3%) than in those with no bone involvement and those with type II hyperostosis (17.0% and 25.8%, respectively, p <0.001). Conversely, *NF2* mutations were less common in the type I hyperostosis group compared to the no bone involvement group and the type II hyperostosis group (17.4% vs. 42.4% vs. 32.3%, p = 0.039). There were no significant differences found for *AKT1* and *KLF4* mutations across the groups.

### Quantitative Bone Density Patterns

Quantitative analysis of CT bone density demonstrated distinct patterns across normal bone and hyperostosis subtypes (Figure 1). In normal bone, HU distributions exhibited a characteristic bimodal pattern, with prominent peaks corresponding to dense cortical bone at the outer and inner edges and a lower-density central distribution consistent with cancellous bone.

Type II hyperostosis preserved this bimodal architecture, with distinct separation between cortical and central ROIs. In contrast, Type I hyperostosis demonstrated a markedly different profile, characterized by a broad, unimodal distribution with substantial overlap between outer, central, and inner ROIs. This pattern reflects loss of normal cortical–cancellous differentiation and more homogeneous bone density throughout the hyperostotic segment.

These quantitative findings corroborated qualitative CT classification, supporting the interpretation that Type I hyperostosis represents architectural disruption, whereas Type II hyperostosis retains native bone structure.

### TRAF7 Mutation Status

To further explore the role of *TRAF7* mutations, we next studied clinical and radiographic differences between *TRAF7*-wildtype and *TRAF7-*mutant tumors across our cohort. Of the 384 meningiomas in our final cohort, 82 (21%) had *TRAF7* mutations (Table 2). Principal component analysis (PCA) of imaging features we collected revealed that *TRAF7-*mutant tumors formed a distinct cluster (Figure 2).

**Figure 2.**
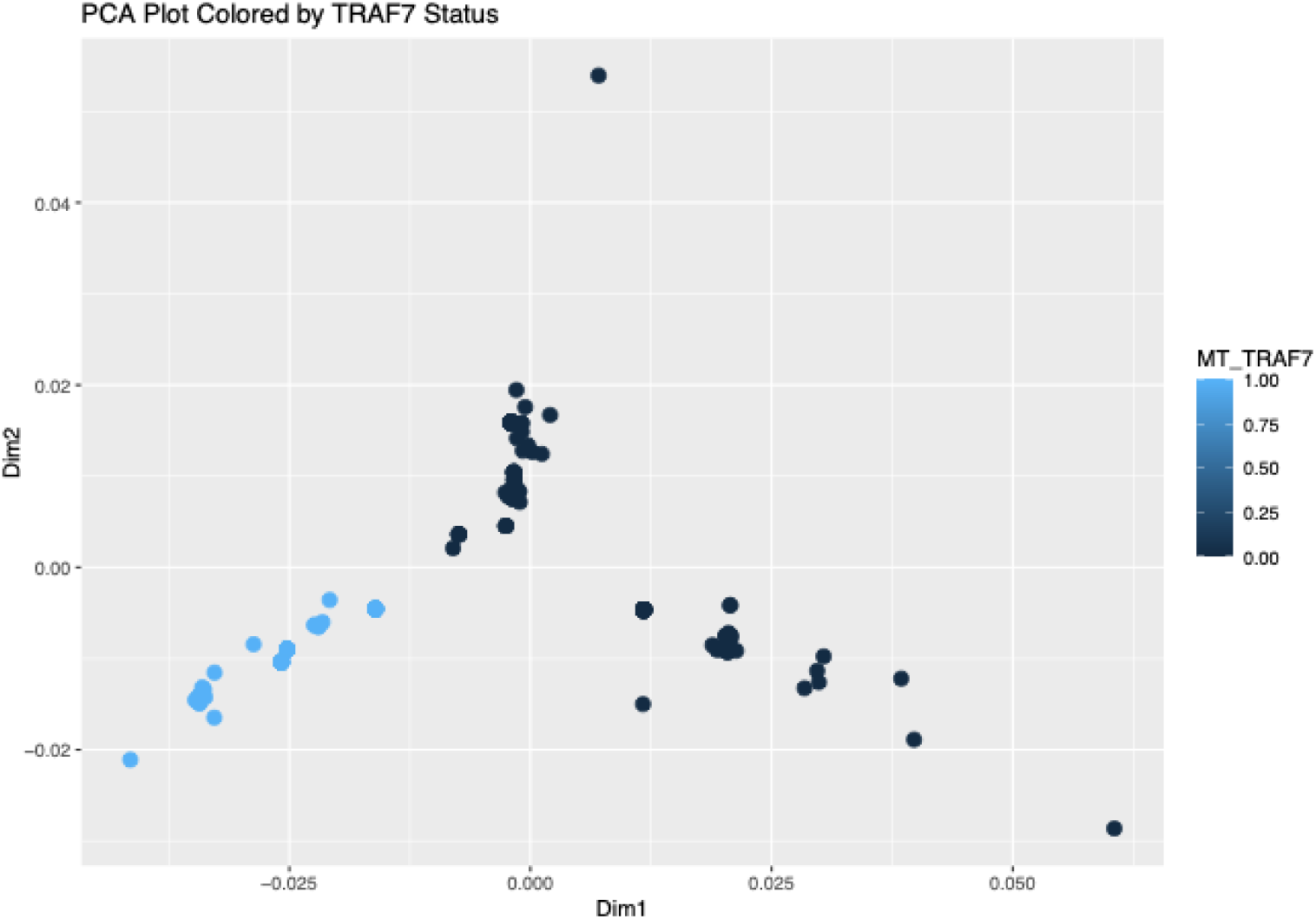
Clustering analysis based on imaging features and *TRAF7* mutation status.

**Table 2.**
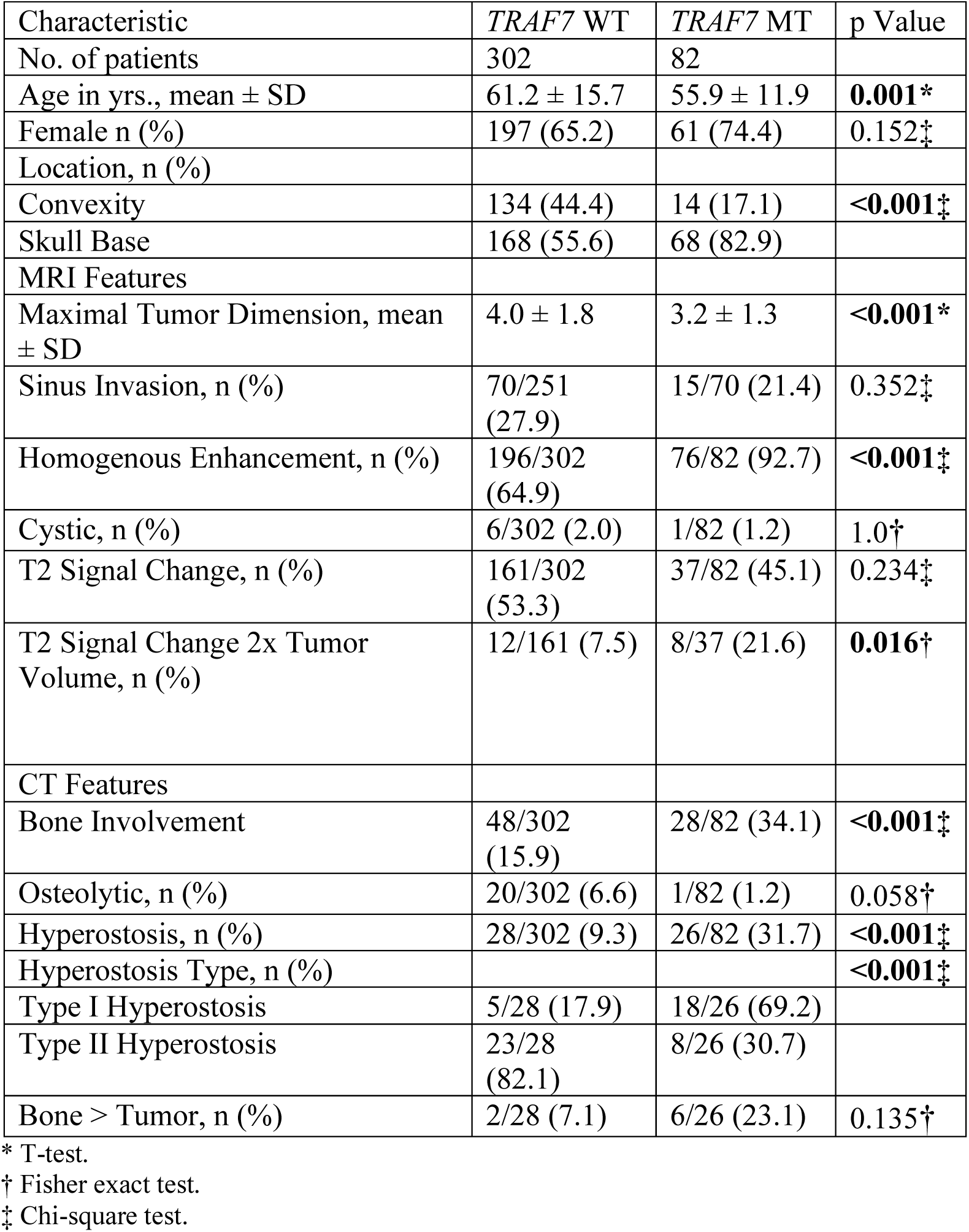
Features by *TRAF7* Mutation.

| Table 2.<br>Features by *TRAF7* Mutation |  |  |  |
| --- | --- | --- | --- |
| Characteristic | *TRAF7* WT | *TRAF7* MT | p Value |
| No. of patients | 302 | 82 |  |
| Age in yrs., mean ± SD | 61.2 ± 15.7 | 55.9 ± 11.9 | **0.001\*** |
| Female n (%) | 197 (65.2) | 61 (74.4) | 0.152‡ |
| Location, n (%) |  |  |  |
| Convexity | 134 (44.4) | 14 (17.1) | **<0.001‡** |
| Skull Base | 168 (55.6) | 68 (82.9) |  |
| MRI Features |  |  |  |
| Maximal Tumor Dimension, mean ± SD | 4.0 ± 1.8 | 3.2 ± 1.3 | **<0.001\*** |
| Sinus Invasion, n (%) | 70/251 (27.9) | 15/70 (21.4) | 0.352‡ |
| Homogenous Enhancement, n (%) | 196/302 (64.9) | 76/82 (92.7) | **<0.001‡** |
| Cystic, n (%) | 6/302 (2.0) | 1/82 (1.2) | 1.0† |
| T2 Signal Change, n (%) | 161/302 (53.3) | 37/82 (45.1) | 0.234‡ |
| T2 Signal Change 2x Tumor Volume, n (%) | 12/161 (7.5) | 8/37 (21.6) | **0.016†** |
| CT Features |  |  |  |
| Bone Involvement | 48/302 (15.9) | 28/82 (34.1) | **<0.001‡** |
| Osteolytic, n (%) | 20/302 (6.6) | 1/82 (1.2) | 0.058† |
| Hyperostosis, n (%) | 28/302 (9.3) | 26/82 (31.7) | **<0.001‡** |
| Hyperostosis Type, n (%) |  |  | **<0.001‡** |
| Type I Hyperostosis | 5/28 (17.9) | 18/26 (69.2) |  |
| Type II Hyperostosis | 23/28 (82.1) | 8/26 (30.7) |  |
| Bone > Tumor, n (%) | 2/28 (7.1) | 6/26 (23.1) | 0.135† |

\* T-test.
† Fisher exact test.
‡ Chi-square test.

Compared to *TRAF7-*wildtype tumors, *TRAF7-*mutant tumors were associated with younger age (55.9 ± 11.9 vs. 61.2 ± 15.7 years, p < 0.001) and were more frequently located in the skull base (82.9% vs. 55.6%, p < 0.001). *TRAF7-*mutant tumors were also found to have smaller maximal intradural tumor dimensions (3.2 ± 1.3 cm vs. 4.0 ± 1.8 cm, p <0.001) and more commonly exhibited homogenous contrast enhancement (92.7% vs. 64.9%, p<0.001). Although the presence of peritumoral edema determined by T2 signal change did now show significant differences amongst these two groups (p = 0.234), extended peritumoral edema, defined as T2 volume hyperintensity that occupies at least twice the tumor volume, was more likely in tumors not carrying a *TRAF7* mutation (21.6% vs. 7.5%, p = 0.016).

Significant differences in features assessed from CT imaging were observed amongst these groups as well. *TRAF7-*mutant tumors had higher rates of bone involvement (34.1% vs 15.9%, p <0.001) and were more likely to present with hyperostosis (31.7% vs. 9.3%, p < 0.001). Among tumors with hyperostosis, most *TRAF7-*mutant tumors exhibited type I hyperostosis (n = 23, 74.2%) while only 5/28 (17.9%) of hyperostotic wild-type tumors showed this pattern (p < 0.001). Of note, the presence of osteolysis was lower among *TRAF7-*mutant tumors compared to wild-type tumors (1.2% vs 6.6%) with this difference approaching statistical significance (p = 0.058).

To evaluate independent predictors of *TRAF7* mutation status, we performed a multivariate logistic regression analysis (Table 3). Variables that were independently associated with *TRAF7* mutations include skull base location (OR 3.11, 95% CI 1.56-6.67, p = 0.002), smaller maximal tumor dimension (OR 0.72, 95% CI 0.59-0.90, p = 0.006), homogenous tumor enhancement (OR 5.57, 95% CI 2.15-17.45, p = 0.001), T2 hyperintensity occupying greater than twice the tumor volume (OR 7.64, 95% CI 2.28-26.51, p = 0.001), and the presence of type I hyperostosis (OR 18.73, 95% CI 3.61-130.92), p = 0.001).

**Table 3.**
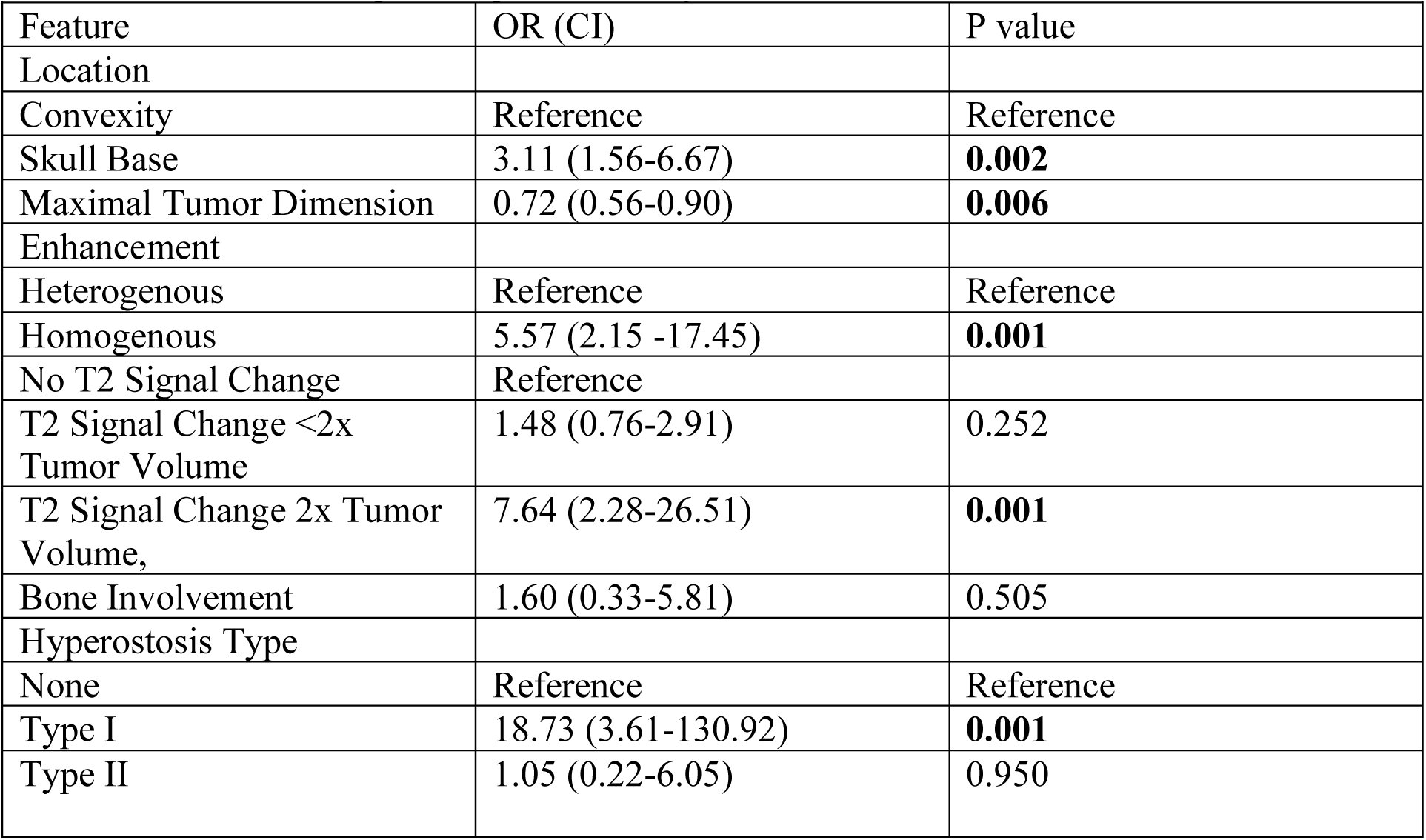
Multivariate logistic regression analysis for TRAF7 mutation status.

To further evaluate the predictive value of these features and to distinguish *TRAF7-*mutants, we employed three machine learning classifiers— Random Forest, GBM, and SVM. GBM achieved the best performance with a test area under the curve (AUC) of 0.85 and a test accuracy of 0.84. Across all three models the most important features included maximal tumor dimension, type I hyperostosis, skull base location, and homogenous enhancement pattern (Figure 3).

**Figure 3.**
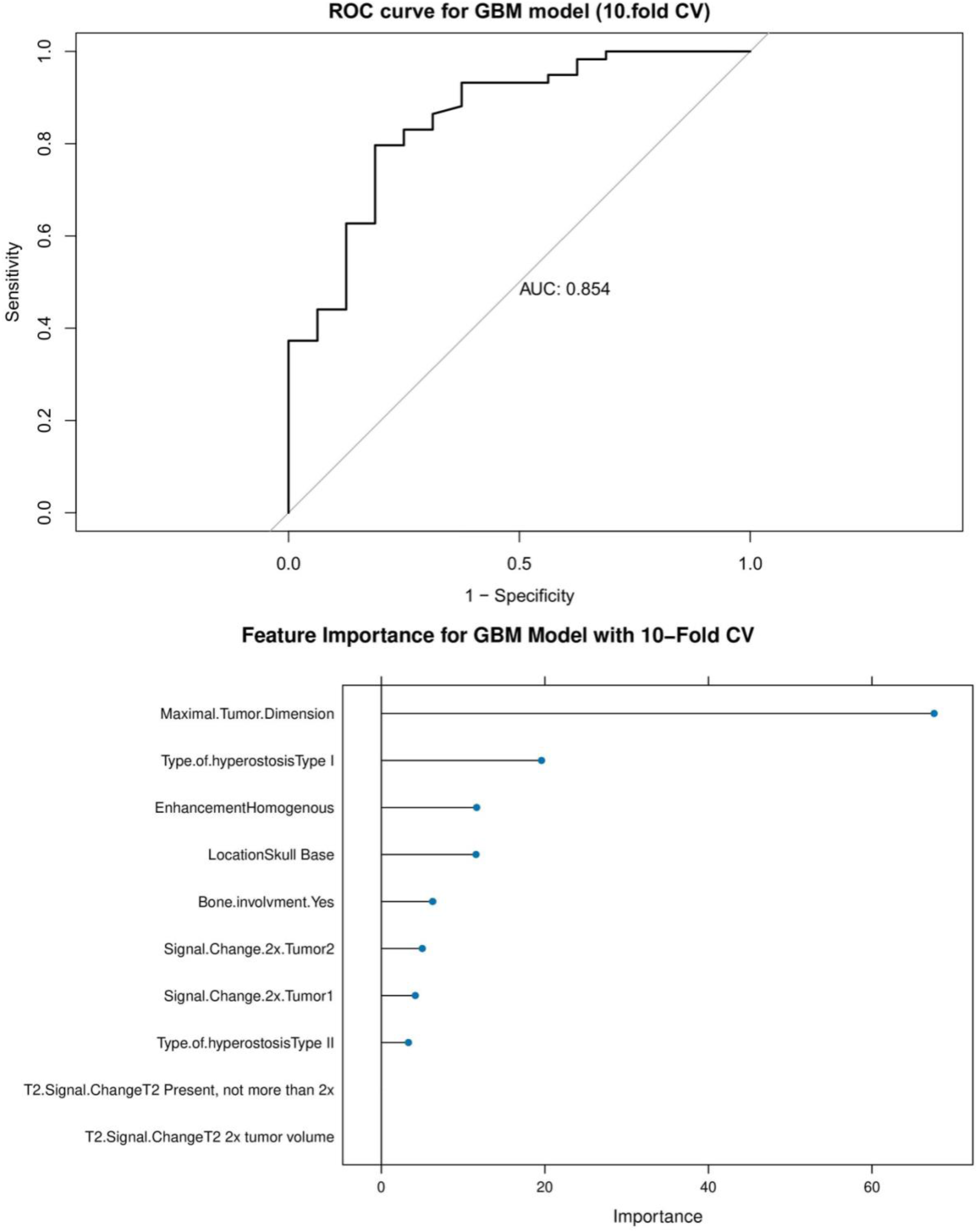
*Top:* Receiver operating characteristic (ROC) curve for the GM model predicting *TRAF7* mutation status. *Bottom*: Feature importance for GBM Model predicting *TRAF7* mutation status using 10-fold cross-validation highlighting the most important contributing features in descending order.

To highlight the defining imaging features and to identify a phenotype highly specific to *TRAF7* mutations, we generated a decision tree classifier. As shown in Figure 4, tumors with type I hyperostosis and an absence of T2 hyperintensity carry a *TRAF7* mutation rate of 92.3%.

**Figure 4.**
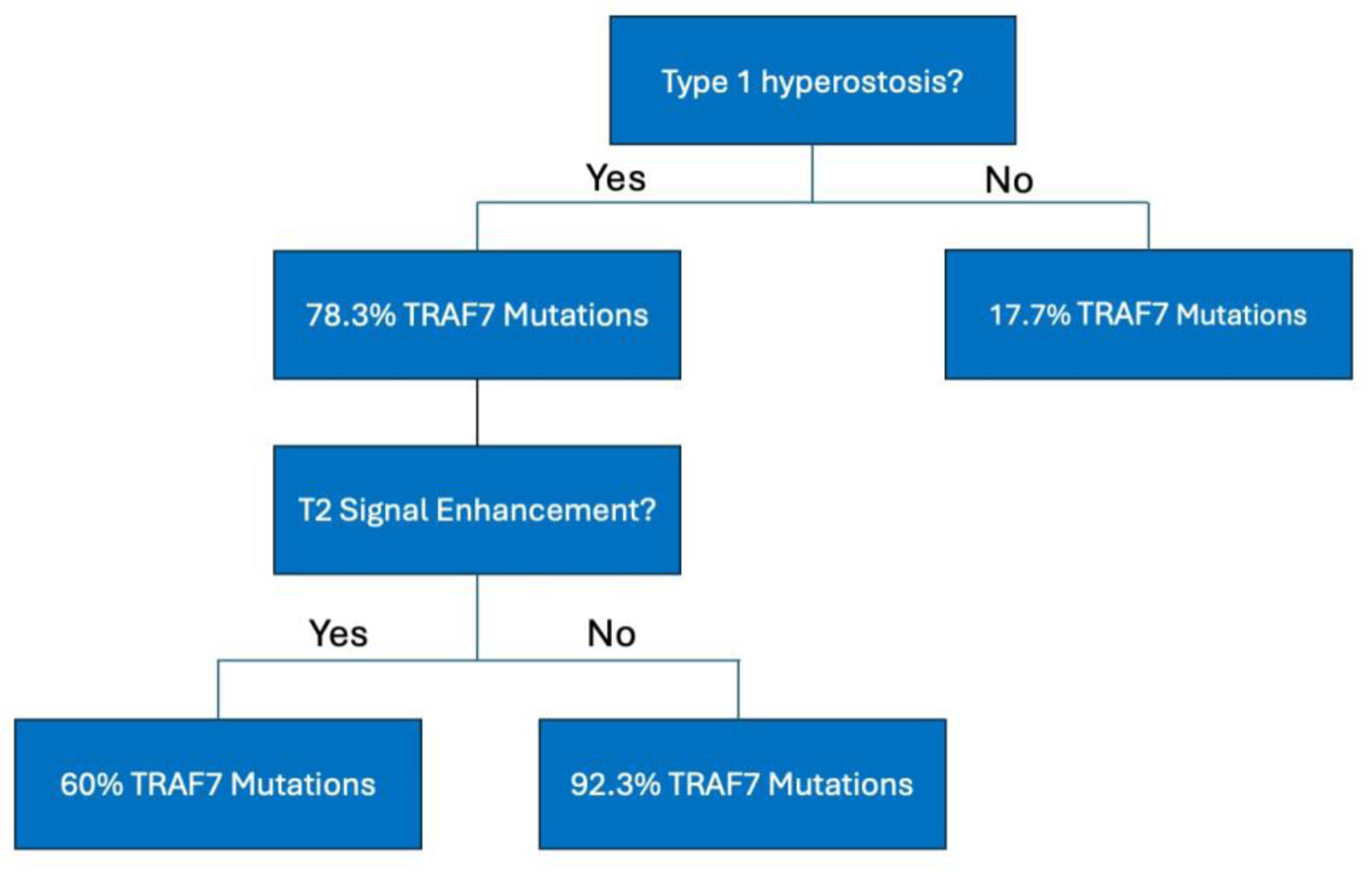
Decision tree illustrating radiographic features that highlight the predictive pathway of *TRAF7* mutation status based on the presence of type of hyperostosis and T2 signal enhancement.

## Discussion

In this study, we investigated the relationship between hyperostosis patterns and associated DNA somatic mutations, particularly *TRAF7*, in patients with intracranial meningiomas. We identified two morphologically distinct subtypes of hyperostosis that are discernible on cross-sectional CT imaging: Type I which is characterized by disruption of cortical-cancellous architecture, and Type II which is defined be the preservation of bone structure. Our results demonstrate that *TRAF7* mutations are strongly associated with Type I hyperostosis, while other commonly observed mutations—such as NF2, AKT1, KLF4— did not show a similar relationship.

### A Distinct TRAF7-Associated Radiographic Phenotype

Beyond hyperostosis, *TRAF7* mutation status was associated with a distinct imaging phenotype: they were more commonly located at the skull base, smaller in maximal dimension, demonstrated homogenous contrast enhancement, and were more likely to demonstrate T2 hyperintensity that occupied at least twice the tumor volume when peritumoral edema was present. Notably, there was a trend that approached significance of osteolysis being less prevalent in *TRAF7-*mutant tumors compared to wild-types. Our multivariate logistic regression confirmed that type I hyperostosis, homogenous enhancement, smaller tumor size, and large-volume T2 hyperintensity were independent predictors of *TRAF7* mutation status. These findings, particularly the skull base location and bony involvement, are consistent with prior studies that have investigated *TRAF7-*mutant meningiomas.^10,13^

The cluster plot from our PCA analysis supports the notion that *TRAF7-*mutant meningiomas exist as a phenotypically distinct subgroup. To further validate our findings, we applied multiple machine learning models to classify *TRAF7* mutation status. Type I hyperostosis and homogenous enhancement remained among the top predictive features in these models. Finally, a decision tree classifier was created to distill these findings into an easily interpretable tool that revealed that tumors with both type I hyperostosis and an absence of T2 hyperintensity had remarkably high rates of *TRAF7* mutations. While no targeted therapies currently exist for *TRAF7-*mutant meningiomas, the ability to noninvasively predict somatic mutations from routine imaging holds clinical value by identifying molecular subtypes that may differ in behavior or response to future therapies. As our understanding of genetic implications in meningiomas grows, such approaches may guide future surgical planning and the integration of radiogenomic markers into personalized care.

### Hyperostosis as a Radiogenomic Signature

Hyperostosis has been a radiographic feature identifiable preoperatively and plays a role in surgical planning and outcomes.^14,15^ While hyperostosis has long been recognized as a radiographic hallmark of meningiomas, its exact mechanisms have remained unclear. Prior studies have suggested either direct tumor invasion into bone or tumor-induced osteoblastic activity as possible mechanisms.⁴, ⁵ Studies involving genetic profiling have highlighted the potential role of various mutations in different biological processes.^10,12^ Furthermore, although tumoral bone involvement has been classified as hyperostotic, osteolytic, or infiltrative in nature, hyperostosis itself has typically been classified in a binary manner.^1,2^

For example, a study by Jin et. al assessed the relationship between genomic subgroups and various extents of bony involvements in sphenoid wing meningiomas.^10^ In this study, bony involvement was defined by no bone invasion, hyperostosis only, tumor invasion only, or both hyperostosis and tumor invasion. The term “tumor invasion only” in this report is hard to interpret since both hyperostotic and osteolytic meningiomas do have bony involvement. *TRAF7* mutations were associated with hyperostosis only, while *NF2* mutations were more frequently seen in cases with tumor invasion. In this context, hyperostosis was studied based only on its presence or absence. This binary classification may overlook potential heterogeneity within hyperostotic tumors that could further correspond with distinct genetic mutations and their potential biological mechanisms.

Preliminary grayscale histogram analysis of hyperostotic bone revealed clear architectural distinctions between subtypes. In Type I cases, density distributions remained consistent across inner, central, and outer regions, mirroring normal bone architecture. In contrast, Type II lesions demonstrated increased density at the margins, suggesting cortical thickening and disruption of trabecular organization.

Overall, our findings suggest that there are distinct morphologic subtypes of hyperostosis and that these subtypes may reflect varying underlying tumor biology and bone remodeling mechanisms. Type I hyperostosis demonstrated a specific association to *TRAF7* while type II did not, suggesting a potential biological interaction between tumor genotype and the bone remodeling process. We offer a new radiologic framework of identifying hyperostosis that may shed light on and align more closely with molecular pathology.

### Limitations

Our study has limitations, particularly related to our data collection processes. The retrospective nature of the study and single-center analysis introduces potential selection bias and limits generalizability. Additionally, patients in this study had to have available genetic panels and imaging which further limits the study’s generalizability. Our classification of hyperostosis subtypes was qualitatively determined on imaging assessment and the consistency of determination of subtype was not formally evaluated. Lastly, the performance of our machine learning models that offered predictive value of mutation status has not been tested in a validation cohort.

### Future Directions

Future work should aim to validate the radiographic subtypes of hyperostosis and associations with known genetic mutations in larger, multi-institutional cohorts. These next steps may inform our understanding of hyperostosis as a phenomenon of meningiomas and may also inform the development of preoperative risk stratification and surgical planning. In addition, current efforts exist to provide a biological link between *TRAF7* mutations and MIH.

## Data Availability

All data produced in the present study are available upon reasonable request to the authors.

